# From Preservation to Regeneration: Stem Cell-Derived Therapies in Machine-Perfused Kidney Transplants – A systematic Review and Meta-analysis

**DOI:** 10.64898/2025.12.22.25342821

**Authors:** Margaux Navez, Elisa Dos Santos Barata, Nathalie Maes, Elena Levtchenko, Fanny Oliveira Arcolino, Perrine Burdeyron, Clara Steichen, Olivier Detry, Nicholas Gilbo, François Jouret

## Abstract

The integration of regenerative medicine into dynamic organ preservation may mitigate ischemia-reperfusion injury in kidney transplantation. This systematic review and meta-analysis evaluated the therapeutic potential of stem cell-based interventions during machine perfusion. Following PRISMA guidelines, PubMed, Embase, and Scopus were searched for experimental studies using stem cells or extracellular vesicles (EVs) during hypothermic or normothermic machine perfusion in animal or discarded human kidneys. Outcomes included renal function, injury biomarkers, inflammation, and histology. Nine studies were included, seven in meta-analysis. Despite heterogeneity in models and protocols, several reported reductions in inflammatory cytokines (e.g., IL-6, IL-1β) and biomarkers (e.g., NGAL) following stem cell or EVs administration. However, meta-analysis showed no significant effects on creatinine clearance (Standardized Means (SMD): 0.00; 95% CI: –0.54 to 0.55), urine output (SMD: 0.54; 95% CI: –0.46 to 1.55), or NGAL (SMD: –1.68; 95% CI: –5.60 to 2.25). Histological protection varied, and stem cell retention was limited. Only one study assessed post-transplant function. While stem cell therapies during perfusion may have immunomodulatory and cytoprotective effects, consistent functional benefits were not observed. Further standardized studies, including transplant models and long-term outcomes, are needed to clarify therapeutic potential and optimize delivery strategies.

## 1. Introduction

Despite advances in kidney transplantation in recent years, the demand for donor organs continues to significantly exceed supply, resulting in prolonged waitlist durations.^1–3^ The ongoing shortage of organs has led to significant efforts to expand the donor pool and increase graft use, driving the development of innovative preservation techniques such as machine perfusion (MP).^4–7^ Although widely used, conventional static cold storage has limitations, such as ischemia-induced cellular damage, suboptimal graft recovery, and high organ discard rates. To overcome these challenges, alternative preservation modalities such as hypothermic and normothermic machine perfusions have been introduced.^8–11^ These dynamic preservation techniques aim to improve organ viability by providing continuous oxygenation. Normothermic perfusion additionally supports active metabolism by delivering nutrients and metabolic substrates, while clearing of waste products throughout the preservation period.^12–14^ Simultaneously, regenerative medicine has become a promising complement to organ preservation, with stem cell-based therapies introducing new strategies to reduce ischemia-reperfusion injury and promote graft repair mechanisms.^15–20^ Specific stem cell populations, notably mesenchymal stromal cells (MSCs), along with their bioactive derivatives such as extracellular vesicles (EVs), display powerful regenerative and immunomodulatory capacities that hold promise for enhancing kidney graft preservation and function.^11,21–23^ Combining these cell-based approaches with advanced perfusion technologies may improve transplant outcomes and bridge the gap between organ supply and demand. This systematic review investigates the therapeutic potential of stem cell-derived products within machine perfusion systems, specifically focusing on their ability to reduce renal ischemia-reperfusion injury and enhance creatinine clearance. The current body of literature on this topic remains highly heterogeneous, notably in terms of perfusion modalities, timing of interventions, analytical methodologies, and types of cellular products. This variability underscores the importance of a comparative analysis of existing studies to elucidate the relative advantages and limitations of each approach. By synthesizing current evidence, our systematic review and meta-analysis aim to provide insights into emerging strategies that could improve kidney graft preservation and post-transplant outcomes.

## 2. Material and Methods

### 2.1. Search Strategy

This systematic review was conducted in accordance with the Preferred Reporting Items for Systematic Reviews and Meta-Analyses (PRISMA) guidelines.^24,25^ A comprehensive literature search was performed in PubMed, Embase, and Scopus on 25 February 2025, using the concepts “stem cell”, “machine perfusion”, and “kidney”, with additional perfusion and stem cell-related terms. The detailed search strategy is presented in **Table S1** and further described in **Supplemental Methods**.

### 2.2. Study Selection and Data extraction

Studies investigating the effect of stem cell-based therapies combined with MP vs. MP alone on graft function or injury in animal models or patients undergoing kidney transplantation were considered. Studies were included if they met the following criteria: (1) published in English or French, (2) original research articles, (3) focusing on kidney transplantation, (4) conducted in a machine perfusion model, and (5) investigating stem cell-based therapies. Studies were excluded if they did not meet these criteria or were unavailable in full-text format. Details are in the **Supplemental Methods**.

### 2.3. Risk of Bias and Quality Assessment

The Systematic review center for laboratory animal experimentation (SYRCLE’s) risk of bias tool for animal studies was used to evaluate study methodologies, with quality assessment of the reported articles based on animal experiments (details in the **Supplemental Methods**).^26^ For studies on human organs, methodological quality was assessed using the National Institutes of Health (NIH) scoring tools.

### 2.4. Statistical Analysis

Renal function was assessed using creatinine clearance during perfusion, after perfusion or after transplantation, and mean urine output during perfusion. Renal blood flow and fractional excretion of sodium were also considered. Renal injury was assessed through Neutrophil Gelatinase-Associated Lipocalin (NGAL) levels. For each outcome, when available, means and standard deviations were extracted for control and treatment groups, along with sample sizes (details in the **Supplemental Methods**). All analyses were performed using the metafor package in R (v4.3.0; R Core Team, Vienna, Austria).

## 3. Results

### 3.1. Study Selection

A total of 1,862 studies were identified through the database search (Pubmed, Embase and Scopus). Aser removal of duplicates identified by Covidence (n=681), 874 studies were screened with titles and abstracts.^27^ During full-text review, 110 of the 119 articles were excluded for the following reasons: five did not involve kidneys, 40 were review articles or conference abstracts, 53 did not involve machine perfusion, and 12 did not involve stem cell–based therapy or derivatives. Thus, nine experimental studies met the predefined inclusion criteria and were included in the qualitative analysis of this systematic review.^15,28–35^ Of these, four studies reported sufficient results for aggregated data analysis on the effect of MSCs treatment during MP on creatine clearance, five on urinary output and three on NGAL levels. The study selection process is summarized in the PRISMA flow diagram **(Figure S1).**

### 3.2. Quality and risk of bias assessment

Risk of bias was mostly “unclear” for animal studies due to insufficient methodological reporting, particularly in relation to selection bias **(Figure S2, Tables S2 and S3)**. Specifically, all studies failed to clearly report on the method of allocation sequence generation and concealment. Baseline group similarity was reported in only one study,^35^ while adjustment for potential confounders remained largely unaddressed. Performance bias was variably reported: random housing of animals is not applicable to most included studies, as they were conducted *ex vivo* rather than involving live animals.^28–31,34^ Nevertheless, only one study explicitly reported blinding of caregivers and investigators, while the others did not provide sufficient information for assessment.^35^ Detection bias was poorly mitigated, with limited reporting on random outcome assessment and assessor blinding. Due to a lack of reporting on the handling and justification of missing data, attrition bias was judged unclear in the majority of studies, except for two studies.^29,35^ Reporting bias was considered low in studies with available protocols or comprehensive outcome reporting.^28–31,35^ Other sources of bias were minimal across studies, with most publications demonstrating freedom from confounding factors such as unit of analysis errors or funding-related influence.^28–31,34,35^ For human studies, the overall quality was better, most often “good” or “not reported” **(Figure S2, Table S4)**.

### 3.3. Study Characteristics

Nine experimental studies met the inclusion criteria, all exploring stem/progenitor cell therapies delivered during *ex vivo* kidney machine perfusion. **Table 1** and **Figure 1** summarize key parameters: species, model design, sample size, and ischemia times.

**Figure 1.**
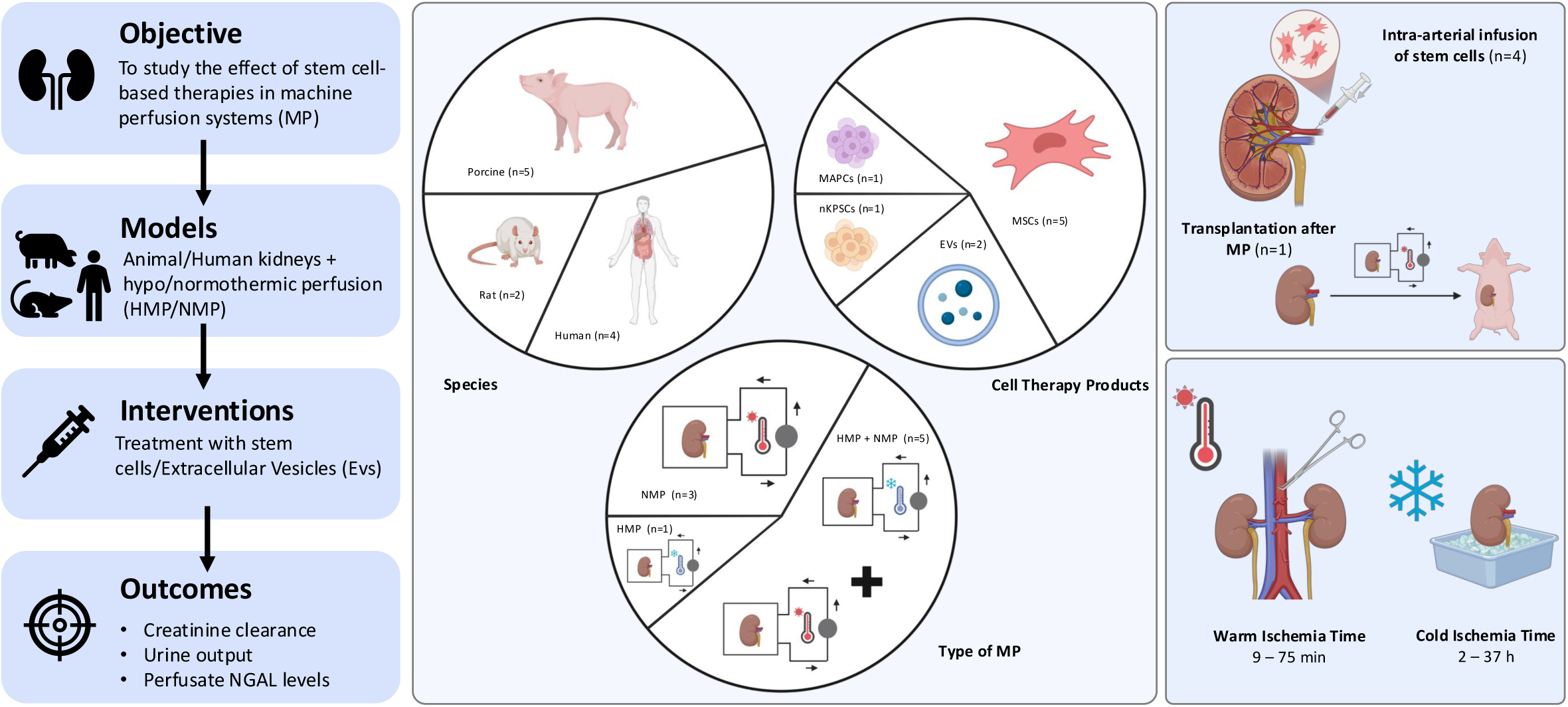
Risk of bias assessment. (A) Risk of bias assessment using SYRCLE’s tool for animal studies; (B) Risk of bias assessment using NIH tools for human studies.

**Table 1.** Overview of the nine included studies summarizing species, groups, model and ischemia times.

| **Study** | **Species** | **N (groups)** | **Model** | **Warm Ischemia Time (WIT)** | **Cold Ischemia Time (CIT)** |
| --- | --- | --- | --- | --- | --- |
| **Lohmann et al, 2021<sup>35</sup>** | Porcine | 4 groups of n=7 | HOPE (14h) + NMP (4h) | 75 min | 120 min |
| **Vallant et al, 2024<sup>34</sup>** | Human<br>Porcine<br>Rodent | 2 groups of n=3<br>2 groups of n=6 | HMP (4h) + NMP (2h) | 25 min | 24h |
| **Rampino et al, 2022<sup>33</sup>** | Human discarded kidneys | 2 groups of n=5 | HOPE (4h) + NMP (4h) | Not indicated | 16-37h |
| **Oliveira Arcolino et al, 2022<sup>32</sup>** | Human discarded kidneys | 2 groups of n=3 | NMP (6h) | 9-18min | 17h |
| **Thompson et al, 2021<sup>15</sup>** | Human discarded kidneys | 2 groups of n =5 | NMP (7h) | Not indicated | 13-36h |
| **Pool et al, 2020<sup>31</sup>** | Porcine | 4 groups of n= 3 | NMP (7h) | 30 min | 3,5-5h |
| **Burdeyron et al, 2024<sup>28</sup>** | Porcine | 3 groups of n=5-6 | HMP (24h) + NMP (5h) | 32 min | / |
| **Pool et al, 2020<sup>30</sup>** | Porcine | 4 groups of n=4 | HMP (2-3h) + NMP (7h) | 20 min | / |
| **Gregorini et al, 2017<sup>29</sup>** | Rat | 3 groups of n=5 | HMP (4h) | 20 min | / |

Four studies employed porcine kidneys^28,30,31,35^, three human discarded kidneys^15,32,33^, one a rat model^29^, and one included all three.^34^ Porcine kidneys were exposed to controlled warm ischemia (20-75 minutes) via renal artery clamping or circulatory arrest (n=6 studies).^28–31,34,35^ In human studies, warm ischemia was uncontrolled, reported in one case as 9-18 minutes.^32^ Most studies used static cold storage, with durations ranging from 2h to 37h (n=6 studies).^15,31–35^ Perfusion protocols varied: typically, kidneys underwent Normothermic Machine perfusion (NMP, at ∼37°C) either alone (n=3 studies)^15,31,32^ or following an initial phase of hypothermic machine perfusion (HMP) or oxygenated hypothermic perfusion (HOPE) (n=5 studies).^28,30,33–35^ A single study investigated HMP alone.^29^ NMP duration ranged from 2 to 7 hours, while HMP/HOPE phases lasted 2-24 hours. Only one study included transplantation and 14-day follow-up.^35^

Cell types investigated were MSCs from adipose tissue (n=1 study),^35^ bone marrow (n=2 studies)^29,34^ or both (n=2 studies)^30,31^ (**Table 2**). One study tested EVs from MSCs as a cell-free therapy alternative.^29^ Others used neonatal kidney stem progenitor cells (nKSPCs)^32^, multipotent adult progenitor cells (MAPCs)^15^ or EVs from porcine urine progenitor cells (UPCs) or MSCs.^28,33^ Most studies compared cell-based therapy to machine perfusion alone (n=8 studies),^15,28–30,32–35^ with additional groups such as porcine vs. human MSCs (n=1 study),^35^ adipose-derived MSCs (A-MSCs) vs. bone marrow-derived MSCs (BM-MSCs) (n=1 study),^30^ or EVs combinations (HMP/EVs + NMP vs. HMP/EVs + NMP/EVs, n=1 study)^28^. Cell dosing varied: two studies used labelled cells at different concentrations (n=2 studies),^31,34^ three used 10 million cells based on safety data;^30,32,35^ two used arbitrary doses (50 million and 3 million).^15,29^ EVs doses were either arbitrarily set (28.5 × 10⁹ EVs)^33^ or based on secretion from cultured cells (6.5 × 10⁹ EVs from 12 million cells).^28^ Administration was mostly via the renal artery during perfusion, often 60 min after NMP initiation;^15,28,30,31,35^ some studies did not specify timing but followed a similar approach.

**Table 2.** Overview of the nine included studies summarizing stem cell type, administrated doses, routes of administration and measured outcomes.

| **Study** | **Cell therapy products** | **Dose** | **Administration** | **Measured Outcomes** |
| --- | --- | --- | --- | --- |
| **Lohmann et al, 2021<sup>35</sup>** | A-pMSCs/ hMSCs | 10 x 10<sup>6</sup> cells | Intra-arterial infusion after 62 min of NMP | Perfusion characteristics, metabolic activity, creatinine clearance, NGAL, IL-6 and IL-10 levels, Y-chromosome detection |
| **Vallant et al, 2024<sup>34</sup>** | BM-rMSCs/hMSCs | 1 x 10<sup>6</sup> cells<br>3 x 10<sup>6</sup> cells<br>5 x 10<sup>6</sup> cells | Not indicated | Perfusion characteristics, creatinine clearance, IL1-β and NGAL levels, localisation of MSCs in kidneys, MHC-1 detection |
| **Rampino et al, 2022<sup>33</sup>** | EVs (MSCs) | 28,5 x10<sup>9</sup> EVs | Not indicated | Renal morphology, ischemic damage, Tubular cell proliferation index and mitochondrial distress, HGF and VEGF tissue levels, apoptosis (caspase 3) |
| **Oliveira Arcolino et al, 2022<sup>32</sup>** | nKSPCs | 10 x 10<sup>6</sup> cells | Intra-arterial infusion 5-10 min before NMP | IL-6, IL-10 and NGAL levels, localisation of stem cells in kidneys, human mixed lymphocyte reaction, cytokines analyses |
| **Thompson et al, 2021<sup>15</sup>** | MAPCs | 50 x 10<sup>6</sup> cells | Infusion after 60 min of NMP | CEUS, EL-8, KIM-1 and NGAL levels, cytokines analyses, MAPCs tracking in kidney tissue |
| **Pool et al, 2020<sup>31</sup>** | BM-MSCs/A-MSCs | 1 x 10<sup>5</sup> cells<br>1 x 10<sup>6</sup> cells<br>1 x 10<sup>7</sup> cells | Intra-arterial infusion after 60 min of NMP | Histology, localisation of stem cells in kidneys, MRI |
| **Burdeyron et al, 2024<sup>28</sup>** | EVs (UPCs) | 6,5 x10<sup>9</sup> EV | Infusion after 60 min of NMP or 30 min of HMP | Perfusion characteristics, biochemical analysis, cytokines analyses, metabolomic, histology and RNAseq |
| **Pool et al, 2020<sup>30</sup>** | BM-MSCs/A-MSCs | 10 x 10<sup>6</sup> cells | Intra-arterial infusion after 60 min of NMP | Perfusion characteristics, cytokines analyses, NGAL and KIM-1 levels, histology |
| **Gregorini et al, 2017<sup>29</sup>** | BM-rMSCs/EVs (MSCs) | 3 x 10<sup>6</sup> cells | Not indicated | Histology, biochemical assays |

Despite model and protocol heterogeneity, studies shared outcome measures: perfusion parameters, renal function (e.g., urine output, creatinine clearance), and cytokine levels. Mechanistic insights were obtained via biochemical, histological, and immunological analyses, including injury biomarkers (e.g., NGAL, IL-6, IL-10), cytokine profiling, advanced imaging (Magnetic Resonance Imaging (MRI), Contract-Enhanced Ultrasound (CEUS)), cell tracking, and transcriptomics.

### 3.4. Composition of the perfusate

**Table 3** summarizes the composition of the perfusate used during HMP and NMP, highlighting substantial heterogeneity in both base solutions and pharmacologic additives. Among the perfusion devices used across the included studies, two employed an adapted pediatric cardiopulmonary bypass system.^15,32^ Four studies utilized a custom-made device based on the LifePort® system (Organ Recovery Systems, Itasca, IL, USA).^28,30,31,35^ One study used the VitaSmart® device from Bridge to Life (London, UK),^33^ another employed the RM3 system (Waters Medical Systems, Rochester, MN, USA),^34^ and the remaining study did not specify the device used.^29^ Most HMP strategies employed University of Wisconsin-based machine perfusion solutions (UW-MPS, Bridge to Like Ltd., Columbia, SC, USA), typically perfused at 4°C and low pressures ranging from 25 to 50 mmHg (n=5 studies).^29,30,33–35^ Exception included the use of KPS-1® (Organ Recovery System, Itasca, IL, USA) for one study.^28^ Notably, HMP perfusates were acellular and not supplemented with pharmacologic additives beyond the base solution. In contrast, NMP protocols consistently incorporated oxygenated, red cell-based perfusates, either with autologous whole blood (n=1 study)^34^ or packed red blood cells (n=7 studies).^15,28,30–33,35^ The perfusate was enriched with additives targeting metabolic support (e.g., glucose, insulin, calcium gluconate, sodium bicarbonate), antimicrobial prophylaxis (e.g., amoxicillin-clavulanate, cefuroxime), and vascular modulation (e.g., verapamil, mannitol). Most protocols operated at physiological temperatures (36–37°C) and perfusion pressures between 70 and 120 mmHg, with oxygenation ensured via carbogen (95% O₂/5% CO₂) at flow rates of 0.2–0.5 L/min. While the core components were similar, differences emerged in the complexity of additives—ranging from basic formulations (e.g., creatinine, glucose, antibiotics) to more elaborate mixtures including multivitamins, Nutriflex® (B. Braun Melsungen AG, Melsungen, Germany) and dexamethasone.^15^ Overall, NMP protocols displayed a convergence toward physiologically active, cell-rich perfusates with targeted metabolic and pharmacologic supplementation.

**Table 3.** Overview of included studies summarizing perfusion settings.

| Study | Device | HMP |  | NMP |  |  |
| --- | --- | --- | --- | --- | --- | --- |
|  |  | Solution | Parameters | Perfusate | Additives | Parameters |
| **Lohmann et al, 2021**<sup>35</sup> | Custom-made device based on LifePort system (Organ Recovery System) | 500 ml cold University of Wisconsin machine perfusion solution (UW-MPS) | • 4-7°C<br>• 0,1 L/min 100% oxygen<br>• 25 mmHg | • 250 ml 5% human albumin solution<br>• 170 ml allogeneic erythrocytes | • Sodium bicarbonate<br>• Glucose<br>• Insulin<br>• Calcium gluconate<br>• Mannitol<br>• Creatinine<br>• Amoxicillin-clavunate<br>• Verapamil | • 37°C<br>• 0,5 L/min carbogen (95% O<sub>2</sub>/ 5% CO<sub>2</sub>)<br>• 70 mmHg |
| **Vallant et al, 2024**<sup>34</sup> | RM3 (Waters Medical Systems) | 500 ml UW-MPS | • 4-6°C<br>• 40-50 mmHg | • 500 ml autologous whole blood<br>• 500 ml of 0.9% saline solution | • Heparin<br>• Cefuroxime<br>• Creatinine | • 36-37°C<br>• 0,2 L/min carbogen (95% O<sub>2</sub>/ 5% CO<sub>2</sub>)<br>• 100 mmHg |
| **Rampino et al, 2022**<sup>33</sup> | VitaSmart device (Bridge to Life) | UW-MPS | • 4°C<br>• 25-30 mmHg | One unit of HBOCs | Non-Reported | • 20-25°C<br>• 60 mmHg |
| **Oliveira Arcolino et al, 2022**<sup>32</sup> | Adapted pediatric cardiopulmonary bypass technology | / |  | Oxygenated red cell-based solution | Non-Reported | • 36°C<br>• 75 mmHg |
| **Thompson et al, 2021**<sup>15</sup> | Adapted pediatric cardiopulmonary bypass technology (Medtronic) | / |  | • 1 unit crossed-matched packed red blood cells<br>• 400 ml Ringer's solution | • Sodium bicarbonate<br>• Heparin<br>• Glucose<br>• Nutriflex infusion<br>• Insulin<br>• Multivitamins<br>• Mannitol<br>• Dexamethasone | • 36.5°C<br>• 75 mmHg |
| **Pool et al, 2020** <sup>31</sup> | Custom-made device based on LifePort system (Organ Recovery System) | / |  | • 350 ml pure red blood cells<br>• 500 ml Williams' Medium E | • Albumin<br>• Amoxicillin-clavunate | • 37°C<br>• 120/80 mmHg |
| **Burdeyron et al, 2024** <sup>28</sup> | LifePort perfusion machine (Organ Recovery System) for HMP / Custom-made device for NMP | 1 L KPS-1 | • 4°C<br>• 35 mmHg | • Autologous blood cells<br>• Human albumin solution | • Sodium bicarbonate<br>• Glucose<br>• Calcium gluconate<br>• Creatinine<br>• Amoxicillin-clavulanic acid<br>• Insulin<br>• Verapamil | • 37°C<br>• 75 mmHg<br>• 0,5 L/min carbogen (95% O<sub>2</sub>/ 5% CO<sub>2</sub>) |
| **Pool et al, 2020** <sup>30</sup> | Kidney Assist Transport device (Organ Assist) and custom-made device based on LifePort system (Organ Recovery System) | UW-MPS | • 4°C<br>• 25 mmHg | • 288 ml pure red blood cells<br>• 100 ml human albumin solution<br>• 300 ml sodium chloride | • Sodium bicarbonate<br>• Glucose<br>• Calcium gluconate<br>• Creatinine<br>• Amoxicillin-clavulanate<br>• Insulin<br>• Verapamil<br>• Augmentin | • 37°C<br>• 110/70 mmHg<br>• 0,5 L/min carbogen (95% O<sub>2</sub>/ 5% CO<sub>2</sub>) |
| **Gregorini et al, 2017** <sup>29</sup> | Non-Reported | UW-MPS | • 4°C | / | / | / |

### 3.5. Functional Outcomes

All studies showed no significant differences in perfusion-related parameters such as renal blood flow, intrarenal resistance, and oxygen consumption between treated and control groups.^15,28,30–35^ Notably, one study reported enhanced cortical and medullary perfusion in MAPCs-treated kidneys after 4 hours of NMP, as measured by CEUS—a non-invasive imaging technique that evaluates microvascular blood flow in real time.^15^ Creatinine clearance during perfusion and fractional sodium excretion were generally similar across groups, though some studies observed modest improvements. For example, one study reported improved creatinine clearance in kidneys perfused with A-MSCs although this difference was not statistically significant.^30^ Two studies observed increased urine output in treated kidneys, ^15,32^ but only one reported a statistically significant difference.^15^ One study reported kidney function measurements over a 14-day period following transplantation, demonstrating significant elevated levels of plasmatic creatinine in control groups compared to the HOPE and NMP+hMSCs groups.^35^ However, others found no significant differences in renal function endpoints after machine perfusion, including urine production.

Meta-analysis on aggregated results concerning creatinine clearance was performed on four studies reporting creatinine clearance during NMP,^28,30,32,35^ and one reporting post-HMP values.^34^ The pooled effect size was not significant (SMD: 0.00; 95% CI: –0.54 to 0.55), indicating no measurable impact. Heterogeneity was low (I² = 0.0%, τ² = 0.00), and Cochran’s Q test did not suggest significant variability between studies (p = 0.81). Results from Lohmann *et al.,* who measured creatinine clearance at day 14 post-transplantation, were consistent with the findings during perfusion (**Figure 2**).^35^ Mean urine output during perfusion was reported in five studies, which were all included in the meta-analysis (**Figure 3**).^15,28,30,32,34^ The pooled analysis yielded an SMD of 0.54 (95% CI: –0.46 to 1.55), with no statistically significant difference between treated and control groups. Heterogeneity was moderate (I² = 66.5%, τ² = 0.84), and Cochran’s Q test indicated significant between-study variability (p = 0.031). In summary, the aggregated results do not demonstrate a consistent link between stem cell-based interventions and functional improvement during perfusion. Meta-analyses with standardized mean differences revealed non-significant pooled effects of stem cell or stem cell-derived product on renal blood flow and fractional sodium excretion with low heterogeneity, as detailed in **Supplementary Results (Figure S3, S4)**.

**Figure 2.**
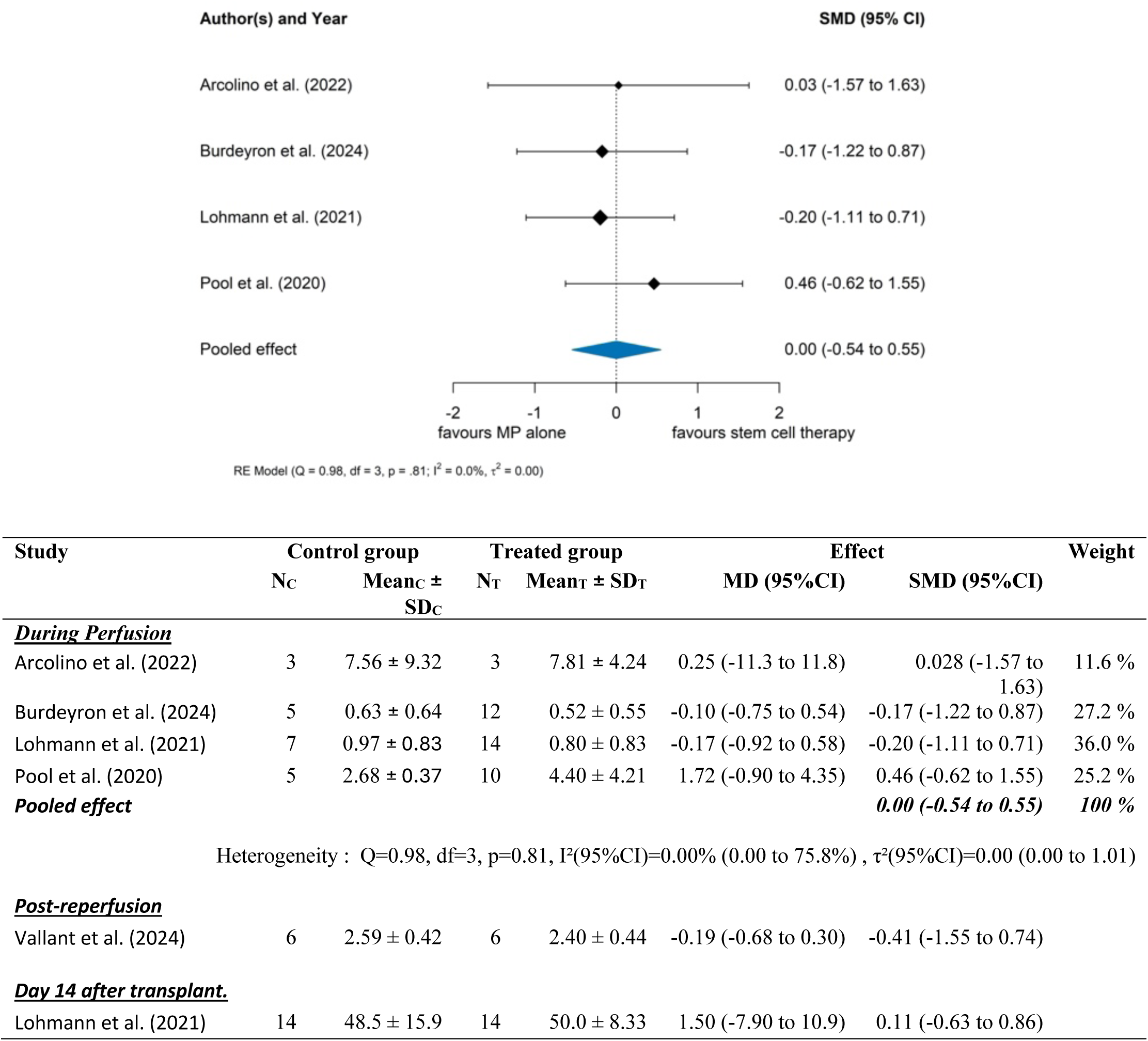
Overview of experimental designs investigating stem cell-based therapies in kidney machine perfusion models. This schematic summarizes key aspects of 9 translational studies evaluating the effects of stem cell–derived therapies in ex vivo kidney perfusion systems. (Les) The primary objective is to assess how stem cells or extracellular vesicles (EVs) influence kidney function using animal (porcine, rat) and human kidneys subjected to hypothermic (HMP) and/or normothermic (NMP) machine perfusion. Reported outcome measures include creatinine clearance, urine output, and perfusate neutrophil gelatinase-associated lipocalin (NGAL) levels. (Center) Studies vary in species used (porcine, rat, human), cell therapy products applied [multipotent adult progenitor cells (MAPCs), kidney progenitor cells (nKPSCs), mesenchymal stromal cells (MSCs), or EVs], and type of machine perfusion (MP) strategy. (Right) Delivery approaches include intra-arterial infusion of stem cells, and organs are subjected to variable warm ischemia times (9–75 min) and cold ischemia times (2–37 h) before perfusion.

**Figure 3.**
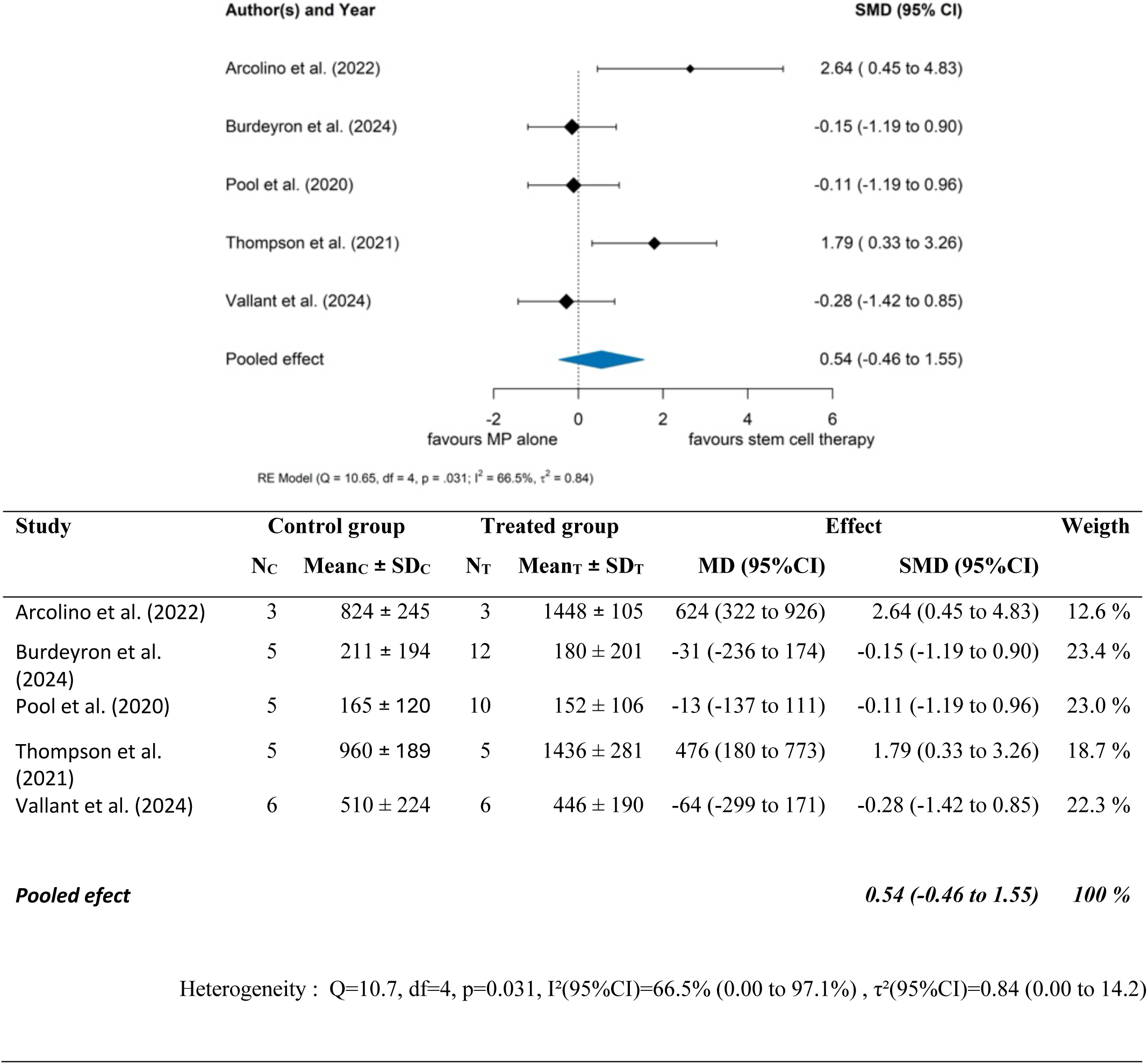
Creatinine Clearance During Perfusion: Meta-Analysis of Standardized Mean Differences. This figure summarizes the results of four studies reporting creatinine clearance during machine perfusion.^28,30,32,35^ The pooled effect size is not statistically significant, with the standardized mean difference (SMD [95% CI]) estimated at 0.00 (–0.54 to 0.55), indicating no effect of stem cell or stem cell-derived product administration on creatinine clearance during perfusion. Heterogeneity across studies was low (I² = 0.0%), with Cochran’s Q test indicating no significant heterogeneity (p = 0.81), and between-study variance τ² = 0.0. Results from Lohmann et al. (2021), assessing creatinine clearance 14 days post-transplantation, and from Vallant et al. (2024), reporting outcomes post-reperfusion are analyzed separately. These findings are consistent with those observed during perfusion.

### 3.6. Injury Markers

Biomarker-based assessment of kidney injury yielded conflicting results. Several studies evaluated NGAL and Kidney Injury Molecule-1 (KIM-1) as primary markers. Three reported decreased NGAL levels in urine or perfusate after treatment with nKSPCs, MAPCs, or MSCs, suggesting potential renoprotection.^15,30,32^ Conversely, other investigations failed to detect significant differences in NGAL or KIM-1 levels between treated and control groups.^28,34,35^ One study noted a progressive rise in plasma NGAL with progressive reduction thereafter although not reaching basal levels over 14 days post-transplantation in all groups, with no intergroup differences.^35^

A meta-analysis of NGAL levels was performed on three studies.^15,28,34^ The study by Lohmann *et al.*, measuring NGAL at day 2 post-transplantation, were analysed separately.^35^ Due to large differences in absolute values between studies, only SMD were used. The pooled SMD was – 1.68 (95% CI: –5.60 to 2.25), indicating no statistically significant difference between treated and control groups (**Figure 4**). Heterogeneity was substantial (I² = 95.5%, τ² = 11.14), and Cochran’s Q test confirmed significant between-study variability (p < 0.001). Post-transplant NGAL results were consistent with perfusion-phase findings. Overall, meta-analysis showed no significant impact of stem cell-based therapies on NGAL levels during or after perfusion.

**Figure 4.**
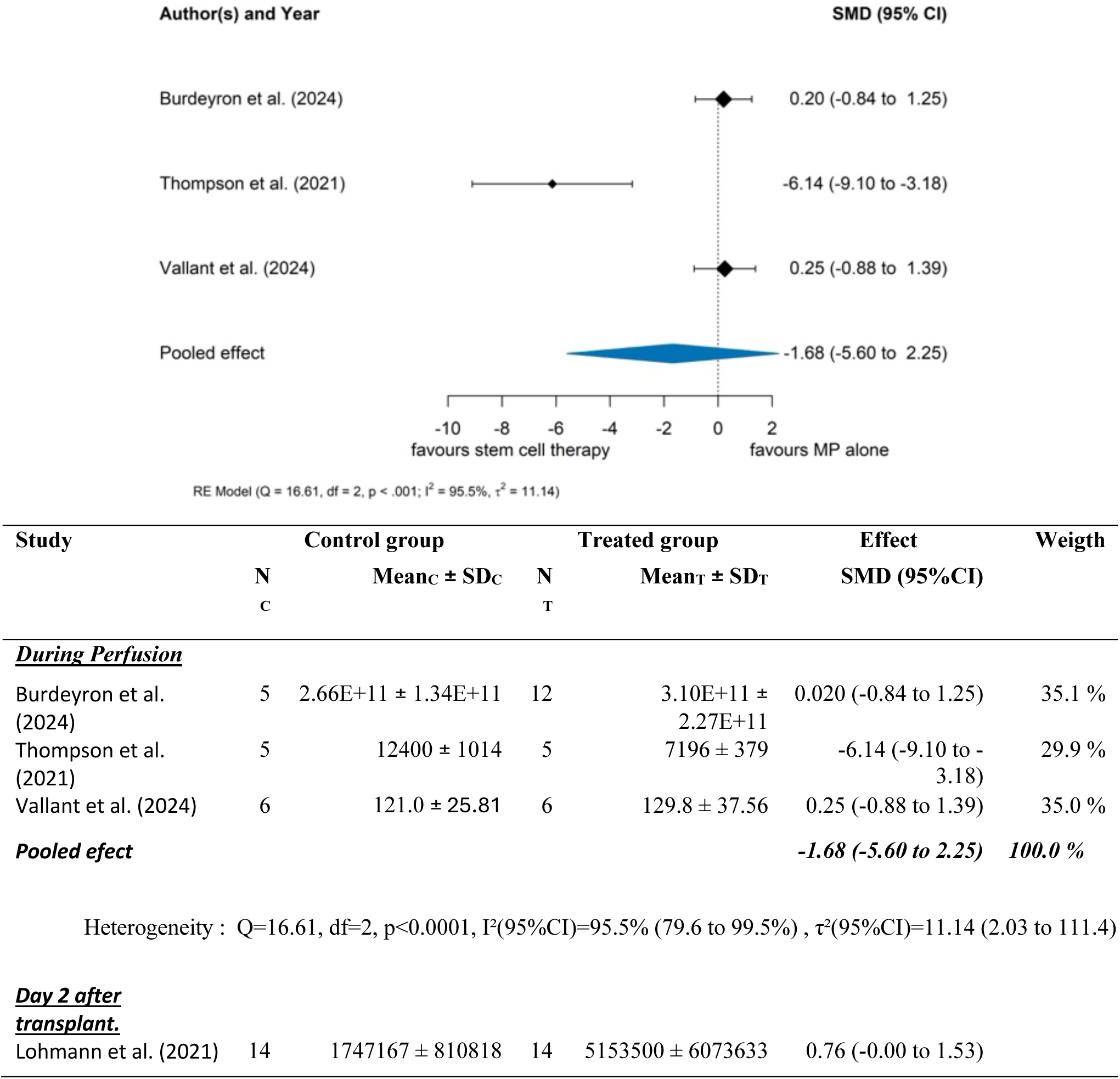
Mean Urine Output During Perfusion: Meta-Analysis of Standardized Mean Differences. This figure presents the pooled analysis of five studies reporting mean urine output during machine perfusion.^15,28,30,34,35^ The overall effect of stem cell or stem cell-derived product administration on urine output is not statistically significant, with a standardized mean difference (SMD [95% CI]) of 0.54 (–0.46 to 1.55). Assessment of heterogeneity revealed moderate variability across studies (I² = 66.5%), with Cochran’s Q test confirming the presence of heterogeneity (p = 0.031) and a between-study variance τ² of 0.84. These results suggest that stem cell-based interventions during machine perfusion do not significantly influence urine output.

**Figure 5.** NGAL Levels During or at the End of Perfusion: Meta-Analysis of Standardized Mean Differences. This figure presents a meta-analysis of three studies reporting NGAL levels during or at the end of machine perfusion.^15,28,34^ NGAL levels reported by Lohmann et al. (2021) at day 2 post-transplantation are analyzed separately.^35^ Due to substantial differences in absolute mean values across studies, only standardized mean differences are presented, not mean differences. The pooled effect is not statistically significant, with an SMD [95% CI] of –1.68 (–5.60 to 2.25), indicating no clear impact of stem cell or stem cell-ssderived product administration on NGAL levels during or at the end of perfusion. Heterogeneity among studies was high (I² = 95.5%), supported by Cochran’s Q test (p < 0.001) and a between-study variance τ² of 11.14. Results for NGAL measured two days post-transplantation are consistent with findings during perfusion.

One study reported reduced ischemic injury in kidneys treated with HOPE plus EVs compared to controls. In the same study, renal caspase-3 expression—an apoptosis marker—was significantly higher in controls than in EV-treated kidneys.^33^ Another study observed no significant difference in leukocyte infiltration between MAPCs-treated and control kidneys, but found reduced flavin mononucleotide (FMN) release, a mitochondrial injury marker, in the treated group.^15^ In HMP settings, another group reported lower levels of tissue injury markers (lactate dehydrogenase, lactate) and oxidative stress, in MSCs- or EVs-treated kidneys compared to controls.^29^ A subsequent study confirmed reduced oxidative stress, as evidenced by decreased glutathione levels, in kidneys treated with both HMP and NMP combined with EVs.^28^

### 3.7. Inflammatory Response

A consistent pattern across studies was the modulation of inflammatory responses following stem cell or EVs administration. Two studies reported a reduction in the pro-inflammatory cytokine interleukin-6 (IL-6) in kidneys subjected to NMP with either nKSPCs or A-MSCs, compared to controls. However, only one of these studies demonstrated a statistically significant decrease.^30^ A separate investigation reported a significant decrease in IL-6 levels two hours post transplantation in the HOPE group relative to controls. However, in the same study, IL-6 secretion increased in all NMP groups to similar levels.^35^ The expression of the pro-inflammatory marker interleukin-1 beta (IL-1β) was significantly decreased in studies using nKSPCs and MAPCs combined with NMP, or HMP/NMP plus EVs, when compared to control groups.^15,28,32^ In contrast, an upregulation of IL-1β was observed in kidneys treated with MSCs in an HMP model, although this increase did not reach statistical significance.^34^ The pro-inflammatory chemokine interleukine-8 (IL-8) levels were found to be significantly reduced in nKSPCs-treated kidneys in one study,^32^ whereas another study reported significant higher IL-8 expression in kidneys treated with BM-MSCs and A-MSCs compared to controls.^30^

Furthermore, one study reported a non-significant reduction in interleukin-18 (IL-18), a pro-inflammatory cytokine, in kidneys treated with HMP or NMP combined with EVs, compared to control groups.^28^ At the mRNA level, IL-10, IL-8, and transforming growth factor-beta (TGF-β)—a cytokine involved in fibrosis and immune regulation—were all significantly reduced in the HOPE group one hour after transplantation, compared to controls.^35^

The anti-inflammatory cytokine interleukin-10 (IL-10) was significantly increased in MAPCs-treated kidneys, but not in other models.^15^ Data from another study showed a significant increase in interleukin-1 receptor antagonist (IL-1RA), an anti-inflammatory mediator, suggesting a potential shift toward an anti-inflammatory profile following treatment with HMP or NMP combined with EVs.^33^ One study also assessed Hepatocyte Growth Factor (HGF) and Vascular Endothelial Growth Factor (VEGF)—key in regeneration and angiogenesis—reporting significantly increased expression in HOPE plus EVs-treated kidneys compared to controls.^33^ Additionally, transcriptomic analyses revealed activation of immunomodulatory genes such as FOXP3, SOCS5, and ISG15, as well as metabolic regulators including Idh2, Ndufs8, and Pdhb, reinforcing the molecular basis for observed immunologic changes.^28,29^

### 3.8. Histological findings

Histological assessment of tissue injury was performed in six out of nine included studies. Across all experimental groups, and independently from the experimental group, common histopathological features included inflammation, tubular atrophy, and varying degrees of fibrosis. Two studies reported significantly reduced tissue damage in kidneys treated with MSCs, EVs, or HOPE combined with EVs compared to controls. Specifically, treated groups exhibited decreased tubular epithelial cell flattening, tubular necrosis, and luminal obstruction compared to control groups.^29,33^ Another study demonstrated improved structural preservation in kidneys treated with HMP/EVs or NMP/EVs, with enhanced tissue integrity observed after 5 hours of NMP. However, histological lesions such as tubular dilation, epithelial detachment, and inflammatory infiltrates were still evident following NMP in this study.^28^

### 3.9. Cell tracking and Localization

Several studies employed tracking techniques to confirm cell delivery and localization within renal tissues. Among them, three studies used fluorescent labelling methods,^15,32,34^ while one study assessed cell retention by quantifying the presence of the Y chromosome in the renal cortex.^35^ MSCs and MAPCs were frequently found in the renal cortex, glomeruli, or peritubular spaces following perfusion.^15,34,35^ One study reported that nKSPCs remained detectable in the renal cortex for up to six hours.^32^ Cell delivery appeared limited in another study.^31^ Only the highest dose (10 million MSCs) led to detectable retention, primarily within glomerular capillaries. However, many of these cells were disintegrated, with fluorescent labelling indicating membrane fragments rather than intact MSCs. Kinetic data further showed that most cells disappeared rapidly from the perfusion circuit, especially when no kidney was connected, suggesting trapping outside the organ. Even with a kidney, only minimal retention occurred after the first passage. No quantification per high-power field was provided, and flow cytometry detected very few MSCs in renal tissue, reinforcing the conclusion that cell engraftment was low and largely ineffective.^31^

## 4. Discussion

This is the first systematic review and meta-analysis assessing the therapeutic potential of combining stem cell-based therapies with dynamic organ preservation to reduce renal ischemia-reperfusion injury and enhance creatinine clearance. Regardless of stem cell source or perfusion temperature, the studies included in this systematic review and meta-analysis did not provide conclusive evidence of a significant impact of stem cell-based intervention during machine perfusion on renal injury and function, although it should be noted that there was considerable methodological variation between studies. Several studies reported decreased levels of pro-inflammatory cytokines and increased anti-inflammatory mediators, suggesting a potential immunomodulatory effect, although a meta-analysis of these outcomes was not possible. Nevertheless, these changes did not translate into a significant functional improvement during perfusion in preclinical studies.

This divergence between effective modulation of inflammation and absence of functional improvement raises several questions about technical limitations and the underlying mechanisms. Although machine perfusion has gained clinical relevance, most clinical studies to date have focused on comparing perfusion modalities (e.g., HMP vs. NMP, with or without oxygen), while the integration of regenerative therapies into these platforms remains largely experimental.^36–39^ Protocols across studies show marked heterogeneity, involving various cell types (e.g., MSCs from bone marrow or adipose tissue, MAPCs, urine-derived progenitors, or EVs) and different ischemic durations, perfusion temperatures, devices, pressures, and perfusate compositions. This variability limits the ability to draw definitive conclusions regarding efficacy or optimal conditions for therapeutic delivery. Moreover, cell dosing remains highly heterogeneous across studies. Few investigations have compared different doses with standardized endpoints, and no consensus has emerged regarding the optimal number of stem cells to use. While higher doses (e.g., 10 million cells) appear necessary for detectable renal retention, even at this level, only a minority of glomeruli show MSCs presence.^31^ Most cells were found confined to glomerular capillaries, with limited migration into the parenchyma. In many cases, fluorescent labeling indicated cell debris rather than intact MSCs, suggesting that cell death occurs shortly after infusion. These observations strongly indicate that cell viability and biodistribution are critical challenges. Quantitative data on how many stem cells reach the renal parenchyma remain insufficient. The rapid decline of MSCs counts from the perfusate, particularly when kidneys are not connected to the circuit, further suggests that many cells are trapped or lost outside the graft. This could explain the absence of a dose-response relationship and reinforce the hypothesis that therapeutic effects are likely mediated via paracrine signaling rather than sustained engraftment.^40–43^ As for the type of therapeutic agent, both stem cells and EVs have been tested, but direct comparisons are rare. Only a few studies assessed different MSCs sources (e.g., bone marrow vs. adipose-derived), and no one provided conclusive evidence favoring one over another. Similarly, comparisons between stem cells and EVs are limited and inconclusive, making it difficult to recommend one strategy over another. Furthermore, the precise mechanism of action remains to be fully elucidated. It is unclear whether the observed effects arise from active secretion by viable cells or from the release of bioactive molecules following cell apoptosis or fragmentation.

A major gap in the current body of evidence is the lack of post-transplantation data. Only one study included in this review used a transplant model and did not report any functional improvement post-transplantation.^35^ Most studies rely on *ex vivo* endpoints, which do not necessarily predict long-term outcomes. Future research should incorporate transplant models with clinically relevant endpoints, including delayed graft function incidence, long-term renal function, and survival.

Finally, the quality of evidence is limited. Risk of bias was often assessed as unclear due to poor reporting of key methodological aspects, such as randomization, blinding, or allocation concealment. Small sample sizes, inconsistent reporting of cell characterization and viability, and variability in perfusion protocols further limit the strength of conclusions that can be drawn. Standardization of delivery protocols, systematic cell tracking, and robust functional assessment in transplant models will be essential to advance the field. To support future study design, **Table S5** summarizes the key parameters that should be considered in experimental protocols investigating stem cell therapies during machine perfusion.

This study has some limitations. Publication bias is possible, as studies with negative or non-significant results are less likely to be published. Outcome heterogeneity may affect the robustness of pooled results. The quality of included studies varied, with some showing methodological limitations. Differences in study design, animal species and donor conditions, and outcome definitions may further limit comparability.

In summary, this systematic review and meta-analysis of preclinical studies on stem cell-based therapies during machine perfusion did not demonstrate significant improvement in renal injury or function during perfusion, regardless of stem cell source or perfusion protocol. However, such interventions might be associated with immunomodulation and increased anti-inflammatory cytokines. Clinical translation will likely depend on overcoming challenges in delivery efficiency, mechanistic understanding, and rigorous validation in transplant models.

## 5. Disclosure

The authors of this manuscript have no conflicts of interest to disclose.

## Data Availability

All data produced in the present study are available upon reasonable request to the authors

## Acknowledgements/Fundings

The authors thank Sandrina Vandenput, reference librarian of the CHU of Liege, for her help and advice in conducting the systematic literature search. This research was funded by the Fonds de la Recherche Scientifique – FNRS (FRIA Fellowship), without involvement in study design, data collection, data-analysis, manuscript preparation and publication decisions. N.G. is supported by a Postdoctoral Clinical Master Specialist Fellowship by the Fund for Scientific Research (FRS-FNRS, 1R00424F). E.L. is supported by the ERC Consolidator Grant NEOGRAFT (CoG-101045467).

## Supplementary Methods

### 1. Search Strategy

All experimental studies investigating the effect of stem cell-based therapies combined with machine perfusion vs. (vehicle) control on graft function, urine production, or tissue lesions in animal models or patients undergoing kidney transplantation were considered for inclusion. Duplicate records were removed, and studies were screened for eligibility using Covidence.^1^ Initially, titles that were clearly irrelevant to the review’s objectives were excluded. The abstracts of the remaining articles were independently evaluated by two reviewers (MN and EDS) to determine their suitability based on the predefined inclusion criteria. Any discrepancies between the two reviewers were resolved through consensus or by contacting a third reviewer (NG) in case of persistent discrepancies. Conference abstracts and other forms of grey literature were not included in this review, as they typically lack the methodological detail required for thorough quality assessment. Furthermore, the examination of reference lists from the included studies did not yield any additional eligible articles beyond those already identified through the systematic search. Subsequently, full-text articles that were retrievable were assessed for final inclusion. The protocol was submitted to PROSPERO on May 13, 2025, prior to commencing the review’s drafting, and is currently awaiting publication.

### 2. Study Selection and Data extraction

Specific outcomes were not pre-defined as inclusion criteria, as we anticipated that the number of studies available on this topic would be limited. All articles meeting the predefined inclusion criteria were considered. No restrictions were applied regarding the publication date to ensure comprehensive coverage of relevant literature. Data extraction was performed in a standardized manner using a prespecified data extraction table in Excel, capturing key study characteristics, including: (1) animal model, (2) sample size, (3) type and duration of perfusion, (4) perfusate composition, temperature, and pressure seconds, (5) stem cells/EVs origin and isolation technique, and (6) timing, duration, administration method, and dose of stem cells/EVs. Outcome parameters varied depending on the specific type of dynamic preservation assessed in each study. Comparison of the therapeutic effects of stem cell-based interventions during machine perfusion was based on the following main reported outcomes: (1) perfusion characteristics (flow, intrarenal resistance), (2) functional outcomes (creatinine clearance and urine output during perfusion), (3) biomarkers of kidney injuries, (4) inflammatory markers, (5) histological findings, and (6) cell/EVs localization or tracking.

### 3. Risk of Bias and Quality Assessment

The Systematic review center for laboratory animal experimentation (SYRCLE’s) risk of bias tool for animal studies derives from the Cochrane Risk of Bias framework but has been specifically refined to capture sources of bias unique to animal intervention studies. It incorporates signaling questions designed to guide systematic and reproducible assessments of bias, thereby enhancing the transparency, rigor, and interpretability of the review findings.^2^

For studies on human organs, methodological quality was assessed using the National Institutes of Health (NIH) scoring tools.

### 4. Statistical Analysis

When only medians and interquartile ranges were reported, means and standard deviations were estimated. When studies reported multiple treatment or control subgroups, these were combined to avoid double-counting. Pooled means and standard deviations were calculated using standard formulas accounting for group sizes and variances. Outcomes were harmonized across studies. To account for different measurement scales, standardized mean differences (SMDs) with 95% confidence intervals (CI) were computed. Forest plots were used to visualize study weights and pooled results. Heterogeneity was assessed using I², τ², and Cochran’s Q statistic. A I² <25% indicated low heterogeneity; >75% indicated high heterogeneity. A p-value <0.05 was considered statistically significant. All analyses were performed using the metafor package in R (v4.3.0; R Core Team, Vienna, Austria).

## Supplementary Tables

**Table S1.**
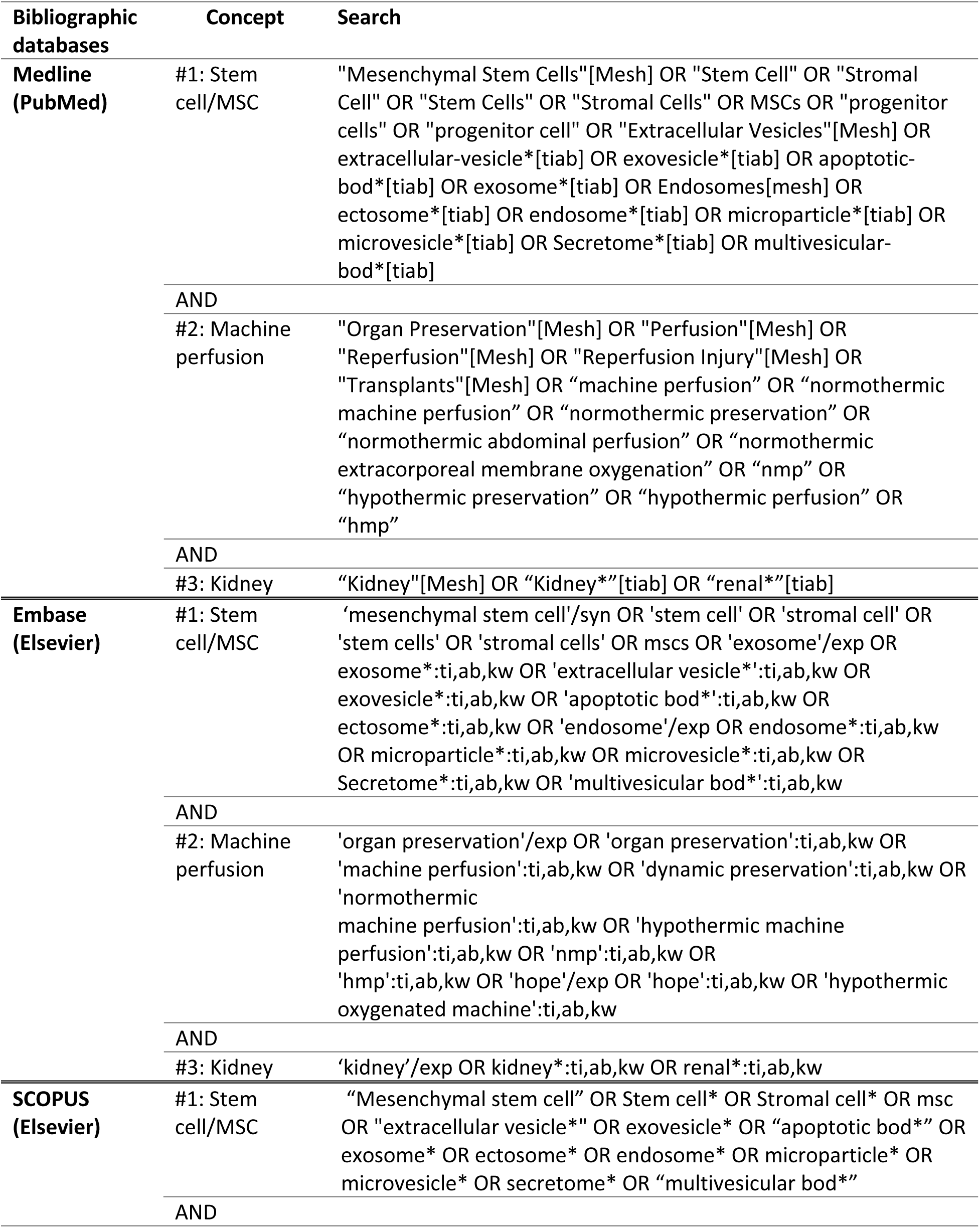
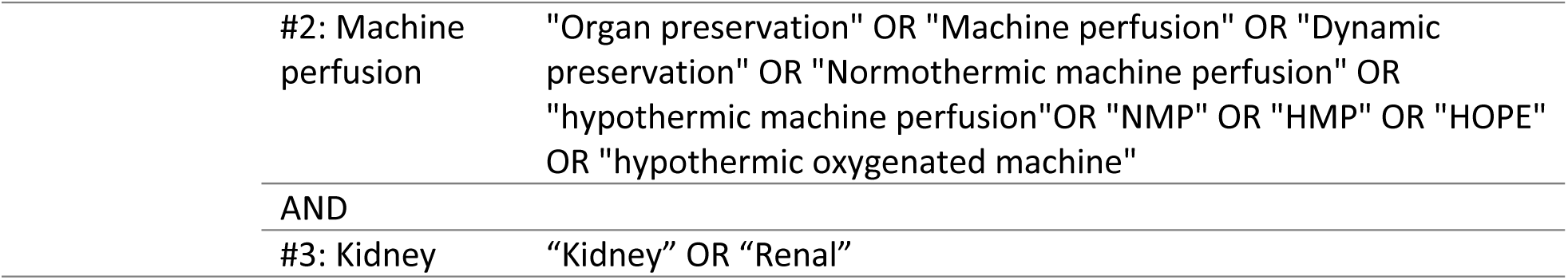
PubMed, Embase and Scopus Search Strategy.

| Bibliographic databases | Concept | Search |
| --- | --- | --- |
| **Medline (PubMed)** | #1: Stem cell/MSC | "Mesenchymal Stem Cells"[Mesh] OR "Stem Cell" OR "Stromal Cell" OR "Stem Cells" OR "Stromal Cells" OR MSCs OR "progenitor cells" OR "progenitor cell" OR "Extracellular Vesicles"[Mesh] OR extracellular-vesicle*[tiab] OR exovesicle*[tiab] OR apoptotic-bod*[tiab] OR exosome*[tiab] OR Endosomes[mesh] OR ectosome*[tiab] OR endosome*[tiab] OR microparticle*[tiab] OR microvesicle*[tiab] OR Secretome*[tiab] OR multivesicular-bod*[tiab] |
|  | AND |  |
|  | #2: Machine perfusion | "Organ Preservation"[Mesh] OR "Perfusion"[Mesh] OR "Reperfusion"[Mesh] OR "Reperfusion Injury"[Mesh] OR "Transplants"[Mesh] OR "machine perfusion" OR "normothermic machine perfusion" OR "normothermic preservation" OR "normothermic abdominal perfusion" OR "normothermic extracorporeal membrane oxygenation" OR "nmp" OR "hypothermic preservation" OR "hypothermic perfusion" OR "hmp" |
|  | AND |  |
|  | #3: Kidney | "Kidney"[Mesh] OR "Kidney*" [tiab] OR "renal*" [tiab] |
| **Embase (Elsevier)** | #1: Stem cell/MSC | 'mesenchymal stem cell'/syn OR 'stem cell' OR 'stromal cell' OR 'stem cells' OR 'stromal cells' OR mscs OR 'exosome'/exp OR exosome*:ti,ab,kw OR 'extracellular vesicle*':ti,ab,kw OR exovesicle*:ti,ab,kw OR 'apoptotic bod*':ti,ab,kw OR ectosome*:ti,ab,kw OR 'endosome'/exp OR endosome*:ti,ab,kw OR microparticle*:ti,ab,kw OR microvesicle*:ti,ab,kw OR Secretome*:ti,ab,kw OR 'multivesicular bod*':ti,ab,kw |
|  | AND |  |
|  | #2: Machine perfusion | 'organ preservation'/exp OR 'organ preservation':ti,ab,kw OR 'machine perfusion':ti,ab,kw OR 'dynamic preservation':ti,ab,kw OR 'normothermic machine perfusion':ti,ab,kw OR 'hypothermic machine perfusion':ti,ab,kw OR 'nmp':ti,ab,kw OR 'hmp':ti,ab,kw OR 'hope'/exp OR 'hope':ti,ab,kw OR 'hypothermic oxygenated machine':ti,ab,kw |
|  | AND |  |
|  | #3: Kidney | 'kidney'/exp OR kidney*:ti,ab,kw OR renal*:ti,ab,kw |
| **SCOPUS (Elsevier)** | #1: Stem cell/MSC | "Mesenchymal stem cell" OR Stem cell* OR Stromal cell* OR msc OR "extracellular vesicle*" OR exovesicle* OR "apoptotic bod*" OR exosome* OR ectosome* OR endosome* OR microparticle* OR microvesicle* OR secretome* OR "multivesicular bod*" |
|  | AND |  |

|  |  |
| --- | --- |
| #2: Machine perfusion | "Organ preservation" OR "Machine perfusion" OR "Dynamic preservation" OR "Normothermic machine perfusion" OR "hypothermic machine perfusion"OR "NMP" OR "HMP" OR "HOPE" OR "hypothermic oxygenated machine" |
| AND |  |
| #3: Kidney | "Kidney" OR "Renal" |

**Table S2:** Detailed Risk of Bias Assessment using SYRCLE’s tool for articles reporting on animal studies.

|  | **References** | Lohmann et al,<br>2021 | Vallant et al,<br>2024 | Pool et al, 2019 | Burdeyron et<br>al, 2024 | Pool et al, 2020 | Gregorini et al,<br>2017 |
| --- | --- | --- | --- | --- | --- | --- | --- |
| Selection<br>bias | Was the allocation sequence adequately generated and applied? | ? | ? | ? | ? | ? | ? |
|  | Were the groups similar at baseline or were they adjusted for confounders in the analysis? | + | ? | ? | ? | ? | ? |
|  | Was the allocation adequately concealed? | ? | ? | ? | ? | ? | ? |
| Performance<br>bias | Were the animals randomly housed during the experiment? | ? | + | + | + | + | + |
|  | Were the caregivers and/or investigators blinded from knowledge which intervention each animal received during the experiment? | + | ? | ? | ? | ? | ? |
| Detection<br>bias | Were animals selected at random for outcome assessment? | ? | ? | ? | + | + | ? |
|  | Was the outcome assessor blinded? | + | ? | ? | ? | ? | ? |
| Attrition<br>bias | Were incomplete outcome data adequately addressed? | + | ? | ? | ? | ? | + |
| Reporting<br>bias | Are reports of the study free of selective outcome reporting? | + | ? | + | + | + | + |
| Other<br>bias | Was the study apparently free of other problems that could result in high risk of bias? | + | + | + | + | + | + |

+, yes = low risk; ?, unclear risk; -, no = high risk; / not relevant or applicable

**Table S3:** SYRCLE’s signaling questions for bias assessment for articles reporting on animal studies.

| Reference | Lohmann et al,<br>2021 | Vallant et al, 2024 | Pool et al, 2019 | Burdeyron et al,<br>2024 | Pool et al, 2020 | Gregorini et al,<br>2017 |
| --- | --- | --- | --- | --- | --- | --- |
| Did the investigators describe a random component in the sequence generation process ? | ? | ? | ? | ? | ? | ? |
| Was the distribution of relevant baseline characteristics balanced for the intervention and control groups? | + | ? | ? | ? | ? | ? |
| If relevant, did the investigators adequately adjust for unequal distribution of some relevant baseline characteristics in the analysis? | / | / | / | / | / | / |
| Was the timing of disease induction adequate? | + | + | + | + | + | + |
| Could the investigator allocating the animals to intervention or control group not foresee assignment due to one of the following or equivalent methods? (*) | ? | ? | ? | ? | ? | ? |
| Did the authors randomly place the cages or animals within the animal room/facility? | ? | ? | ? | ? | ? | ? |
| Is it unlikely that the outcome or the outcome measurement was influenced by not randomly housing the animals? | ? | + | + | + | + | + |
| Was blinding of caregivers and investigators ensured, and was it unlikely that their blinding could have been broken? | + | ? | ? | ? | ? | ? |
| Did the investigators randomly pick an animal during outcome assessment, or did they use a random component in the sequence generation for outcome assessment? | ? | ? | ? | + | + | ? |
| Was blinding of the outcome assessor ensured, and was it unlikely that blinding could have been broken? | + | ? | ? | ? | ? | ? |
| Was the outcome assessor not blinded, but do review authors judge that the outcome is not likely to be influenced by lack of blinding? | / | / | / | / | / | / |
| Were all animals included in the analysis? | + | + | + | + | + | + |
| Were the reasons for missing outcome data unlikely to be related to true outcome? (e.g., technical failure) | + | ? | ? | ? | ? | + |
| Are missing outcome data balanced in numbers across intervention groups, with similar reasons for missing data across groups? | + | ? | ? | ? | ? | ? |
| Are missing outcome data imputed using appropriate methods? | ? | ? | ? | ? | ? | ? |
| Was the study protocol available and were all of the study's pre-specified primary and secondary outcomes reported in the current manuscript? | ? | ? | ? | ? | ? | ? |
| Was the study protocol not available, but was it clear that the published report included all expected outcomes? | + | ? | + | + | + | + |
| Was the study free of contamination (pooling drugs)? | + | + | + | + | + | + |
| Was the study free of inappropriate influence of funders? | + | + | + | ? | + | + |
| Was the study free of unit of analysis errors? | + | + | + | + | + | + |
| Were design-specific risks of bias absent? | + | + | + | + | + | + |
| Were new animals added to the control and experimental groups to replace drop-outs from the original population? | + | + | + | + | + | + |

+, yes = low risk; ?, unclear risk; -, no = high risk; / not relevant or applicable

**Table S4:** Quality assessment pf studies including human kidneys according to the NIH quality assessment score.

|  | References | Vallant et al, 2024 | Rampino et al, 2022 | Oliveira Arcolino et al, 2022 | Thompson et al, 2020 |
| --- | --- | --- | --- | --- | --- |
| Described as randomized | Was the study described as randomized, a randomized trial, a randomized clinical trial, or an RCT? | NR | NR | NR | NR |
|  | Adequate randomization: was the method of randomization adequate (i.e., use of randomly generated assignment)? | NR | + | NR | NR |
| Treatment allocation-two interrelated pieces | Allocation concealment: was the treatment allocation concealed (so that assignments could not be predicted)? | NR | NR | NR | NR |
| Blinding | Were study participants and providers blinded to treatment group assignment? | NR | NR | NR | NR |
|  | Were the people assessing the outcomes blinded to the participants group assignments? | NR | NR | NR | NR |
| Similarity of groups at baseline | Were the groups similar at baseline on important characteristics that could affect outcomes (e.g., demographics, risk factors, co-morbid conditions)? | - | + | - | + |
| Dropout | Was the overall drop-out rate from the study at endpoint 20% or lower of the number allocated to treatment? | + | + | + | + |
|  | Was the differential drop-out rate (between treatment groups) at endpoint 15 percentage points or lower? | + | + | + | + |
| Adherence | Was there high adherence to the intervention protocols for each treatment group? | - | + | + | + |
| Avoid other interventions | Were other interventions avoided or similar in the groups (e.g., similar background treatments)? | - | + | + | + |
| Outcome measures assessment | Were outcomes assessed using valid and reliable measures, implemented consistently across all study participants? | + | + | + | + |
| Power calculation | Did the authors report that sample size was sufficiently large to be able to detect a difference in the main outcome between groups with at least 80% power? | NR | NR | NR | NR |
| Prespecified outcomes | Were outcomes reported or subgroups analyzed prespecified (i.e., identified before analyses were conducted)? | + | + | + | + |
| Intention-to-treat analysis | Were all randomized participants analyzed in the group to which they were originally assigned, i.e., did they use an intention-to-treat analysis? | NR | NR | NR | NR |

+, yes = good; ?, Other\* = fair; -, no=poor
\*Other: CD, cannot determine; NR, not reported

**Table S5:** Recommended Parameters for Designing Future Experimental Protocols Involving Cell Therapy During Organ Perfusion.

| **Category** | **Parameter to Consider** | **Recommendation / Rationale** |
| --- | --- | --- |
| **Injury model** | Warm Ischemia Time (WIT) / Cold Ischemia Time (CIT) | Precisely document to allow for meaningful comparisons across studies |
| **Perfusion System** | Hypothermic (HMP) vs. Normothermic (NMP) | Clearly specify the mode; cell effects may vary depending on temperature |
| **Perfusate Composition** | Type (blood-based, acellular, with supplements) | Standardize as much as possible to reduce biological variability |
| **Cell Type** | MSCs, endothelial cells, EV | Report cell source, phenotype, viability, and characterization methods |
| **Delivery Method** | Bolus, continuous infusion, intra-arterial administration | Choose a consistent and traceable delivery route; include systematic tracking (e.g., labeling) |
| **Perfusion Duration** | Total perfusion time & time of cell delivery | Provide exact timing to ensure reproducibility and interpretation of effects |
| **Cell Tracking** | Labeling, imaging, histology, Spatial transcriptomics, AI-based tissues analysis | Include standardized tracking to evaluate cell fate and localization |
| **Functional Assessment** | Post-perfusion transplant model | Use robust functional endpoints (e.g., histology, biomarkers, graft survival) |
| **Study Design** | Randomization, blinding, allocation concealment | Ensure methodological rigor to reduce risk of bias |
| **Sample Size** | A priori calculation if feasible | Optimize statistical power to support valid conclusions |

## Supplementary Figures

**Figure S1.**
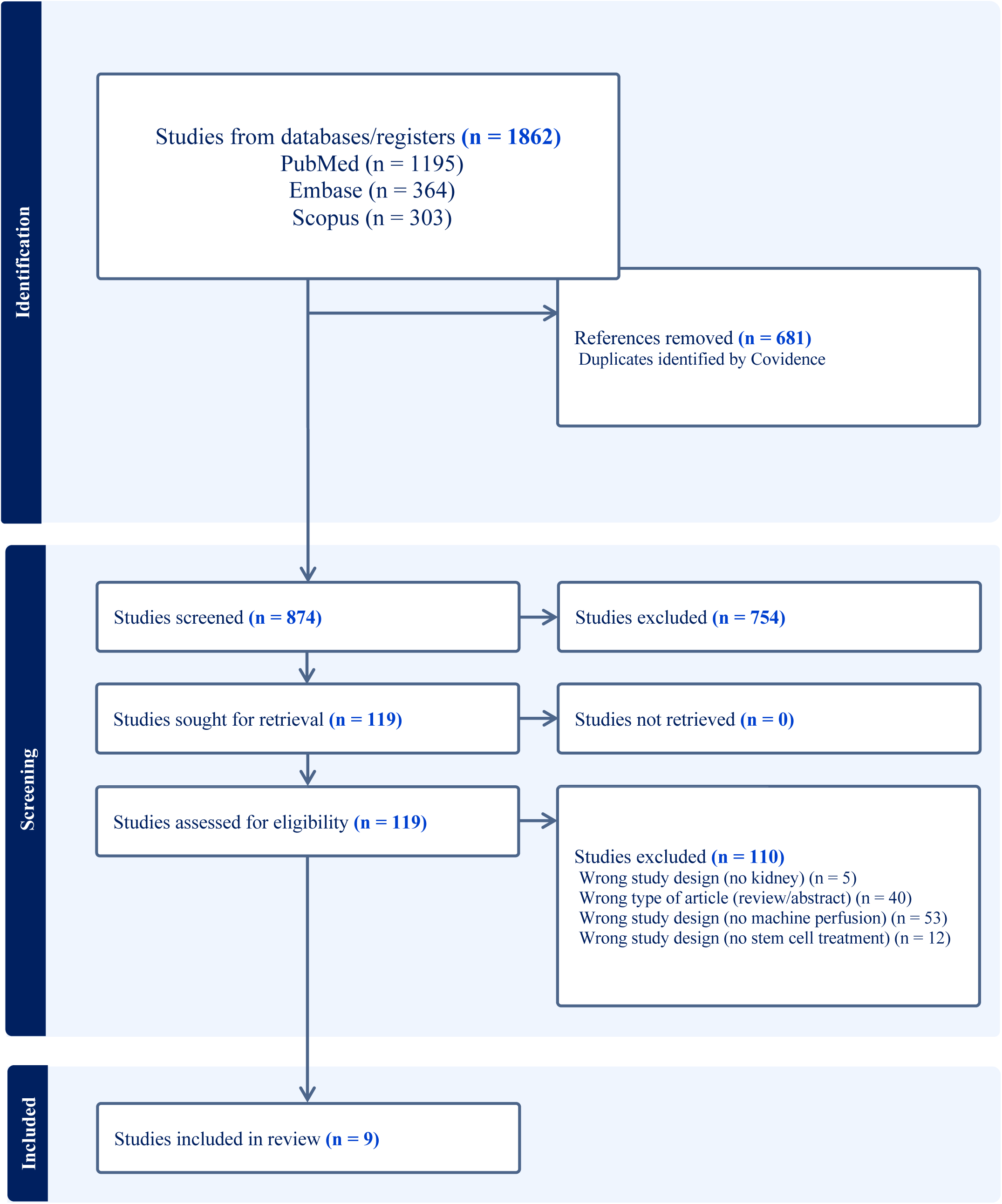
PRISMA Flow Diagram

**Figure S2.**
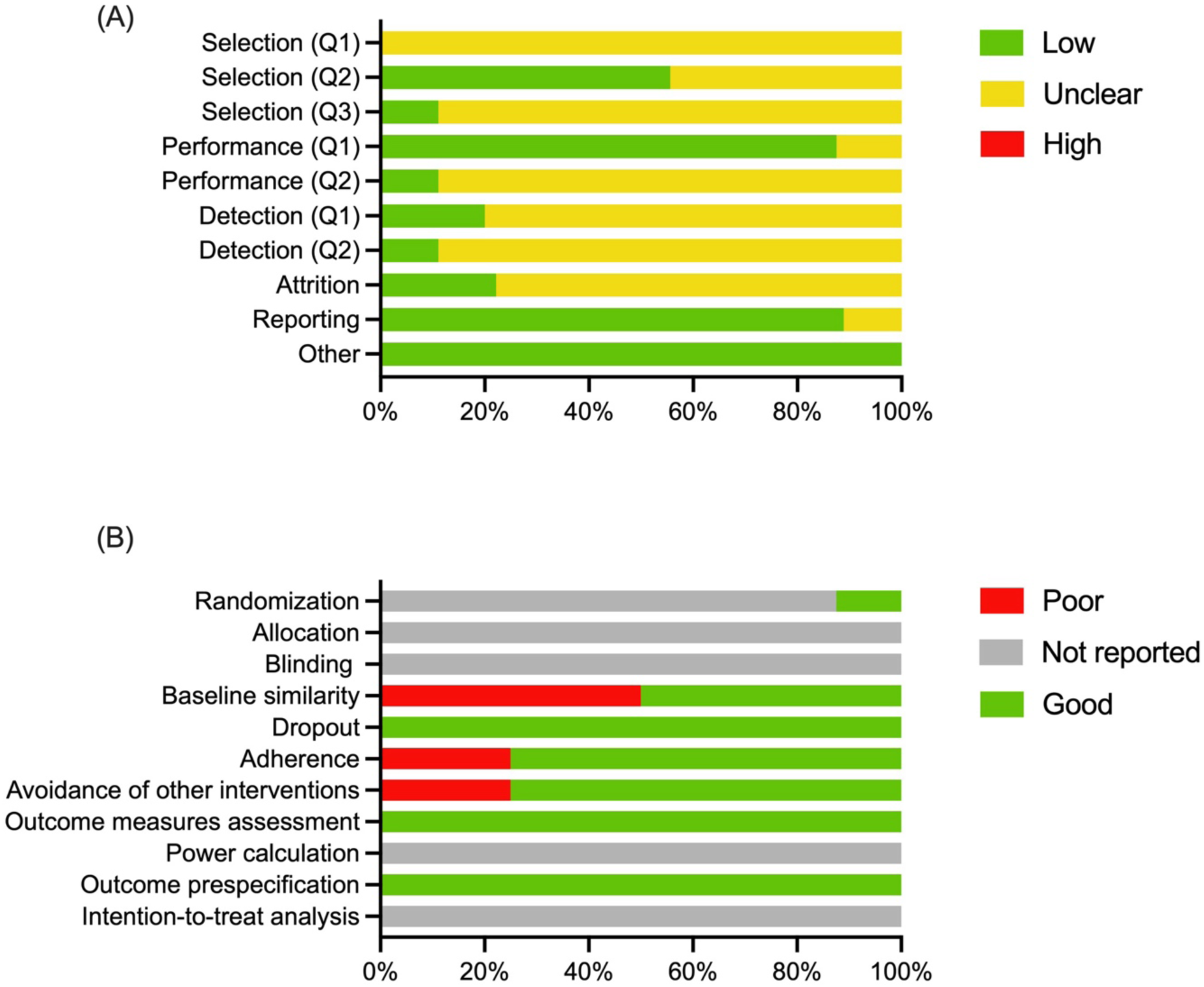
Risk of bias assessment. (A) Risk of bias assessment using SYRCLE’s tool for animal studies; (B) Risk of bias assessment using NIH tools for human studies.

**Figure S3.**
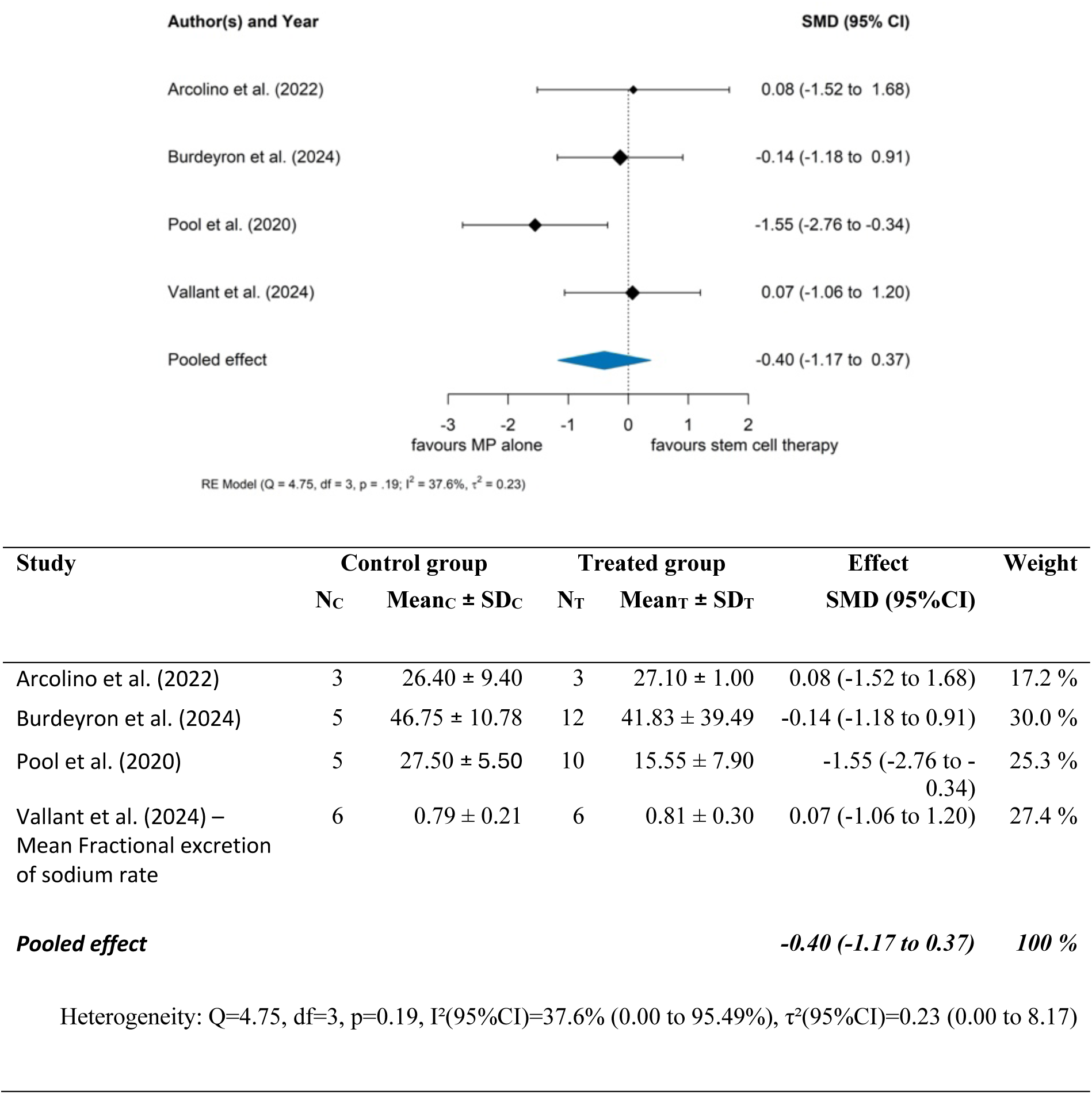
Fractional excretion of sodium (%) during perfusion: Meta Analysis of Standardized Mean Differences. This figure summarizes the meta-analysis of four studies evaluating fractional excretion of sodium during machine perfusion. Due to differences in units of measurement in Vallant et al. (2024), only standardized mean differences are reported. The pooled effect is not statistically significant, with an SMD [95% CI] of –0.40 (–1.17 to 0.37), indicating no measurable impact of stem cell or stem cell-derived product administration on fractional excretion of sodium during perfusion. Heterogeneity across studies was low to moderate (I² = 37.6%), with no significant heterogeneity detected by Cochran’s Q test (p = 0.19), and between-study variance τ² = 0.23.

**Figure S4.**
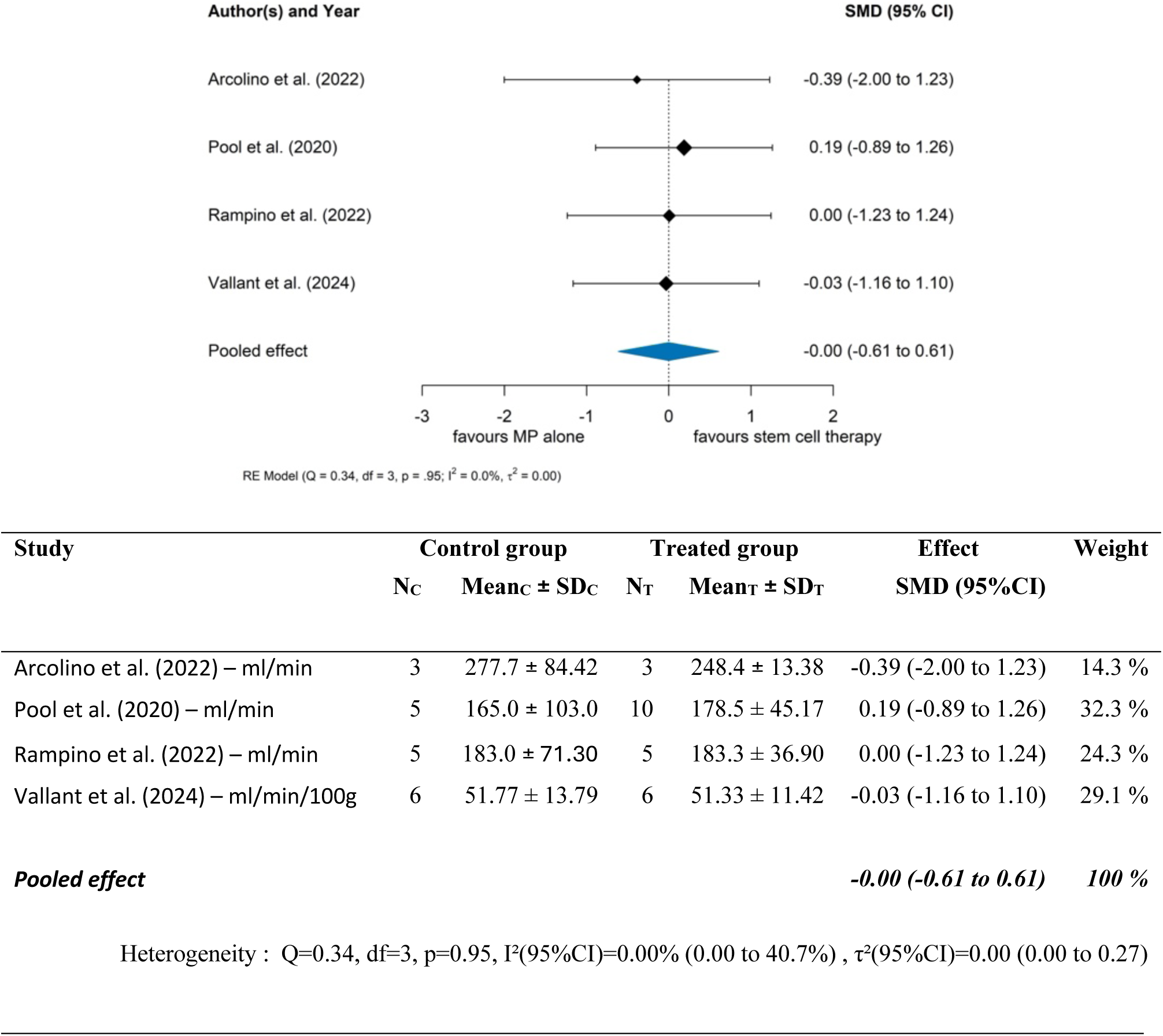
Renal Blood Flow during or at the end of the perfusion: Meta Analysis of Standardized Mean Differences. This figure presents the meta-analysis of four studies assessing renal blood flow during or at the end of machine perfusion. Due to differences in measurement units in the Vallant et al. (2024) study, only standardized mean differences are reported. The pooled effect is not statistically significant, with an SMD [95% CI] of –0.00 (–0.61 to 0.61), indicating no observable effect of stem cell or stem cell-derived product administration on renal blood flow during perfusion. Heterogeneity across studies was minimal (I² = 0.0%), with Cochran’s Q test rejecting heterogeneity (p = 0.95), and between-study variance τ² = 0.0.

## References

1. Chaudhry D, Chaudhry A, Peracha J, Sharif A. Survival for waitlisted kidney failure patients receiving transplantation versus remaining on waiting list: systematic review and meta-analysis. BMJ. 2022;376:e068769. doi:10.1136/bmj-2021-068769

2. Yang J, Endo Y, Munir MM, et al. Waitlist Time, Age, and Social Vulnerability: Impact on the Survival Benefit of Deceased Donor Kidney Transplantation Versus Long-term Dialysis Among Patients With End-stage Renal Disease. Transplantation. 2025;109(1):e64–e74. doi:10.1097/TP.0000000000005125

3. Wolfe RA, Ashby VB, Milford EL, et al. Comparison of Mortality in All Patients on Dialysis, Patients on Dialysis Awaiting Transplantation, and Recipients of a First Cadaveric Transplant. N Engl J Med. 1999;341(23):1725–1730.

4. Ozgur OS, Namsrai BE, Pruett TL, et al. Current practice and novel approaches in organ preservation. Front Transplant. 2023;2. doi:10.3389/frtra.2023.1156845

5. Hasjim BJ, Sanders JM, Alexander M, Redfield RR, Ichii H. Perfusion Techniques in Kidney Allograft Preservation to Reduce Ischemic Reperfusion Injury: A Systematic Review and Meta-Analysis. Antioxidants. 2024;13(6):642.

6. Weissenbacher A, Vrakas G, Nasralla D, Ceresa CDL. The future of organ perfusion and re-conditioning. Transpl Int. 2019;32(6):586–597. doi:10.1111/tri.13441

7. Abbas SH, Friend PJ. Principles and current status of abdominal organ preservation for transplantation. Surgery in Practice and Science. 2020;3:100020. doi:10.1016/j.sipas.2020.100020

8. Almeida S, Snyder W, Shah M, et al. Revolutionizing deceased donor transplantation: How new approaches to machine perfusion broadens the horizon for organ donation. Transplantation Reports. 2024;9(3):100160. doi:10.1016/j.tpr.2024.100160

9. Hosgood SA, Brown RJ, Nicholson ML. Advances in Kidney Preservation Techniques and Their Application in Clinical Practice. Transplantation. 2021;105(11):e202–e214.

10. Jing L, Yao L, Zhao M, Peng L ping, Liu M. Organ preservation: from the past to the future. Acta Pharmacol Sin. 2018;39(5):845–857. doi:10.1038/aps.2017.182

11. Navez M, Gilbo N, Vandermeulen M, et al. Simultaneous Ex Situ Normothermic Perfusion of Paired Kidneys in Pigs. Artificial Organs. Published online May 2, 2025. doi:10.1111/aor.15016

12. Hosgood SA, Callaghan CJ, Wilson CH, et al. Normothermic machine perfusion versus static cold storage in donation after circulatory death kidney transplantation: a randomized controlled trial. Nat Med. 2023;29(6):1511–1519.

13. Radi G, Fallani G, Germinario G, Busutti M, Manna GL, Ravaioli M. HYPOTHERMIC PERFUSION OF THE KIDNEY: FROM RESEARCH TO CLINICAL PRACTICE. European Journal of Transplantation. Published online March 21, 2023:79–91. doi:10.57603/EJT-011

14. Mazilescu LI, Urbanellis P, Kim SJ, et al. Normothermic Ex Vivo Kidney Perfusion for Human Kidney Transplantation: First North American Results. Transplantation. 2022;106(9):1852.

15. Thompson ER, Bates L, Ibrahim IK, et al. Novel delivery of cellular therapy to reduce ischemia reperfusion injury in kidney transplantation. Am J Transplant. 2021;21(4):1402–1414.

16. Hoogduijn MJ, Montserrat N, van der Laan LJW, et al. The emergence of regenerative medicine in organ transplantation: 1st European Cell Therapy and Organ Regeneration Section meeting. Transpl Int. 2020;33(8):833–840. doi:10.1111/tri.13608

17. Lathan R, Ghita R, Clancy MJ. Stem Cells to Modulate IR: a Regenerative Medicine-Based Approach to Organ Preservation. Curr Transpl Rep. 2019;6(2):146–154. doi:10.1007/s40472-019-00240-7

18. Erpicum P, Weekers L, Detry O, et al. Infusion of third-party mesenchymal stromal cells after kidney transplantation: a phase I-II, open-label, clinical study. Kidney International. 2019;95(3):693–707.

19. Vandermeulen M, Erpicum P, Weekers L, et al. Mesenchymal Stromal Cells in Solid Organ Transplantation. Transplantation. 2020;104(5):923–936.

20. Detry O, Jouret F, Vandermeulen M, et al. [Mesenchymal stromal cells and organ transplantation]. Rev Med Liege. 2014;69 Spec No:53-56.

21. Luijmes SH, Verstegen MMA, Hoogduijn MJ, et al. The current status of stem cell- - based therapies during ex vivo graft perfusion: An integrated review of four organs. 2022;(July):1–17. doi:10.1111/ajt.17161

22. Rigo F, De Stefano N, Navarro-Tableros V, et al. Extracellular Vesicles from Human Liver Stem Cells Reduce Injury in an Ex Vivo Normothermic Hypoxic Rat Liver Perfusion Model. Transplantation. 2018;102(5):e205–e210. doi:10.1097/TP.0000000000002123

23. Blondeel J, Gilbo N, De Bondt S, Monbaliu D. Stem cell Derived Extracellular Vesicles to Alleviate ischemia-reperfusion Injury of Transplantable Organs. A Systematic Review. Stem Cell Rev Rep. 2023;19(7):2225–2250. doi:10.1007/s12015-023-10573-7

24. Page MJ, Moher D, Bossuyt PM, et al. PRISMA 2020 explanation and elaboration: updated guidance and exemplars for reporting systematic reviews. BMJ. Published online March 29, 2021:n160. doi:10.1136/bmj.n160

25. Moher D, Liberati A, Tetzlaff J, Altman DG, for the PRISMA Group. Preferred reporting items for systematic reviews and meta-analyses: the PRISMA statement. BMJ. 2009;339(jul21 1):b2535–b2535. doi:10.1136/bmj.b2535

26. Hooijmans CR, Rovers MM, De Vries RB, Leenaars M, Ritskes-Hoitinga M, Langendam MW. SYRCLE’s risk of bias tool for animal studies. BMC Med Res Methodol. 2014;14(1). doi:10.1186/1471-2288-14-43

27. Covidence - Better systematic review management. Covidence. Accessed August 7, 2025. https://www.covidence.org/

28. Burdeyron P, Giraud S, Lepoittevin M, et al. Dynamic conditioning of porcine kidney grafts with extracellular vesicles derived from urine progenitor cells: A proof-of-concept study. Clin Transl Med. 2024;14(12):e70095. doi:10.1002/ctm2.70095

29. Gregorini M, Corradetti V, Pattonieri EF, et al. Perfusion of isolated rat kidney with Mesenchymal Stromal Cells/Extracellular Vesicles prevents ischaemic injury. Journal of Cellular and Molecular Medicine. 2017;21(12):3381–3393. doi:10.1111/jcmm.13249

30. Pool MBF, Vos J, Eijken M, et al. Treating Ischemically Damaged Porcine Kidneys with Human Bone Marrow- And Adipose Tissue-Derived Mesenchymal Stromal Cells during Ex Vivo Normothermic Machine Perfusion. Stem Cells and Development. 2020;29(20):1320–1330. doi:10.1089/scd.2020.0024

31. Pool M, Eertman T, Parraga JS, et al. Infusing mesenchymal stromal cells into porcine kidneys during normothermic machine perfusion: Intact MSCs can be traced and localised to Glomeruli. International Journal of Molecular Sciences. 2019;20(14). doi:10.3390/ijms20143607

32. Arcolino FO, Hosgood S, Akalay S, et al. De novo SIX2 activation in human kidneys treated with neonatal kidney stem/progenitor cells. American Journal of Transplantation. Published online 2022. doi:10.1111/ajt.17164

33. Rampino T, Gregorini M, Germinario G, et al. Extracellular Vesicles Derived from Mesenchymal Stromal Cells Delivered during Hypothermic Oxygenated Machine Perfusion Repair Ischemic/Reperfusion Damage of Kidneys from Extended Criteria Donors. Biology (Basel). 2022;11(3). doi:10.3390/biology11030350

34. Vallant N, Wolfhagen N, Sandhu B, Hamaoui K, Papalois V. Delivery of Mesenchymal Stem Cells during Hypothermic Machine Perfusion in a Translational Kidney Perfusion Study. IJMS. 2024;25(9):5038. doi:10.3390/ijms25095038

35. Lohmann S, Pool MBF, Rozenberg KM, et al. Mesenchymal stromal cell treatment of donor kidneys during ex vivo normothermic machine perfusion: A porcine renal autotransplantation study. American Journal of Transplantation. 2021;21(7):2348–2359. doi:10.1111/ajt.16473

36. Tingle SJ, Figueiredo RS, Moir JAG, Goodfellow M, Talbot D, Wilson CH. Machine perfusion preservation versus static cold storage for deceased donor kidney transplantation. Cochrane Database of Systematic Reviews. 2019;2019(3). doi:10.1002/14651858.CD011671.pub2

37. Tingle SJ, Thompson ER, Figueiredo RS, et al. Normothermic and hypothermic machine perfusion preservation versus static cold storage for deceased donor kidney transplantation. Cochrane Database Syst Rev. 2024;7(7):CD011671. doi:10.1002/14651858.CD011671.pub3

38. Peng P, Ding Z, He Y, Zhang J, Wang X, Yang Z. Hypothermic Machine Perfusion Versus Static Cold Storage in Deceased Donor Kidney Transplantation: A Systematic Review and Meta-Analysis of Randomized Controlled Trials. Artif Organs. 2019;43(5):478–489. doi:10.1111/aor.13364

39. Rijkse E, Bouari S, Kimenai HJAN, et al. Additional Normothermic Machine Perfusion Versus Hypothermic Machine Perfusion in Suboptimal Donor Kidney Transplantation: Protocol of a Randomized, Controlled, Open-Label Trial. Int J Surg Protoc. 2021;25(1):227–237.

40. Maacha S, Sidahmed H, Jacob S, et al. Paracrine Mechanisms of Mesenchymal Stromal Cells in Angiogenesis. Stem Cells International. 2020;2020(1):4356359. doi:10.1155/2020/4356359

41. Alvites R, Branquinho M, Sousa AC, Lopes B, Sousa P, Maurício AC. Mesenchymal Stem/Stromal Cells and Their Paracrine Activity—Immunomodulation Mechanisms and How to Influence the Therapeutic Potential. Pharmaceutics. 2022;14(2):381. doi:10.3390/pharmaceutics14020381

42. Linero I, Chaparro O. Paracrine Effect of Mesenchymal Stem Cells Derived from Human Adipose Tissue in Bone Regeneration. PLoS One. 2014;9(9):e107001. doi:10.1371/journal.pone.0107001

43. Gilbo N, Blondeel J, Pirenne J, Romagnoli R, Camussi G, Monbaliu D. Organ Repair and Regeneration During Ex Situ Dynamic Preservation: The Future is Nano. Transpl Int. 2023;36:11947. doi:10.3389/ti.2023.11947

## References

1. Covidence - Better systematic review management. Covidence. Accessed August 7, 2025. https://www.covidence.org/

2. Hooijmans CR, Rovers MM, De Vries RB, Leenaars M, Ritskes-Hoitinga M, Langendam MW. SYRCLE’s risk of bias tool for animal studies. BMC Med Res Methodol. 2014;14(1). doi:10.1186/1471-2288-14-43

